# Physician Deaths from Corona Virus Disease (COVID-19)

**DOI:** 10.1101/2020.04.05.20054494

**Authors:** Edsel B. Ing, Alis (Qinyuan) Xu, Ali Salimi, Nurhan Torun

**Affiliations:** University of Toronto Ophthalmology, Staff, Toronto, Ontario Canada; University of British Columbia, Medical Student, Vancouver, BC, Canada; McGill University, Medical Student, Montreal, QC, Canada; Harvard Beth Israel Deaconness, Ophthalmology Chief, Boston, MA,USA

**Keywords:** COVID-19, coronavirus, physician, death

## Abstract

**OBJECTIVE:** The COVID-19 pandemic has caused much morbidity and mortality to patients but also health care providers. We tabulated the cases of physician deaths from COVID-19 associated with front-line work in hopes of mitigating future events.

**METHOD:** On April 5, 2020, Google internet search was performed using the keywords “doctor” “physician” “death” “COVID” “COVID-19” and “coronavirus” in English and Farsi, and in Chinese using the Baidu search engine.

**RESULTS:** We found 198 physician deaths from COVID-19, but complete details were missing for 49 individuals. The average age of the physicians that died was 63.4 years (range 28 to 90 years) and the median age was 66 years of age. Ninety percent of the deceased physicians were male (175/194). General practitioners and emergency room doctors (78/192), respirologists (5/192), internal medicine specialists (11/192) and anesthesiologists (6/192) comprised 52% of those dying. Two percent of the deceased were epidemiologists (4/192), 2% were infectious disease specialists (4/192), 5% were dentists (9/192), 4% were ENT (8/192), and 4% were ophthalmologists (7/192). The countries with the most reported physician deaths were Italy (79/198), Iran (43/198), China (16/198), Philippines (14/198), United States (9/192) and Indonesia (7/192).

**CONCLUSION:** Physicians from all specialties may die from COVID, and these deaths will likely increase as the pandemic progresses. Lack of personal protective equipment was cited as a common cause of death. Consideration should be made to exclude older physicians from front-line work.

## INTRODUCTION

The coronavirus disease 2019 (COVID-19) pandemic has infected at least 1.2 million individuals worldwide with an overall 5.5% risk of death as of April 5, 2020. ^(1)^ Physicians caring for COVID-19 infected patients are at high risk of contagion and possible mortality. To quantify and mitigate this risk, the underlying characteristics of physician deaths from COVID infection are investigated.

## METHOD

No research ethics board approval was required for this publicly available information.

On April 5, 2020, a Google internet search was performed using the keywords “doctor”, “physician”, “death”, “COVID”, “COVID-19” and “corona” in English, and repeated in Farsi. An additional search was performed on Baidu (the Chinese version of Google) using the equivalent Chinese characters. (see Appendix) An additional PubMed search was performed using the English search terms.

Physicians who died after contracting COVID-19 from patient care activities or interactions with medical colleagues were included. The doctor’s age, practice focus, gender, country and the date of the report were recorded. Physicians who died from a cardiac event in the line of duty, or over 90 years of age were also noted, but not included in the final database. Data were analyzed using Stata 15.1.

## RESULTS

On PubMed search on April 5, 2020, no articles on physician deaths and COVID-19 were found. Our internet search found 193 physician deaths related to COVID-19 infection due to front-line work or interactions with work colleagues. The date of the reports ranged from February 7, 2020, to April 5, 2020. Complete age details were only available on 149 physicians. The Italian National Federation of Medicine Surgery and Odontology ^(2)^ had a complete list of physician deaths in the country from COVID-19. The deaths in other countries required compilation from numerous web articles. Two physicians caring for COVID-19 patients were thought to die from cardiac compromise or exhaustion but were not diagnosed to have COVID infection, and were excluded from the database. ^(3) (4)^ A doctor that died from COVID infection acquired during travel rather than frontline-work or work colleagues was also excluded. ^(5)^

Physicians older than 90 years of age were excluded from this study. The age details were missing for 49 individuals. The average age of the physicians that died was 63.5 years (range 28 to 90 years) and the median age was 66.0 years. Physicians 57 years of age or older accounted for three-quarters of COVID-related deaths.

Ninety percent (175/194) of the deceased physicians were male. General practitioners and emergency room doctors (78/192), respirologists (5/192), internal medicine specialists (11/192) and anesthesiologists (6/192) comprised 52% of those dying. Two percent of the deceased were epidemiologists (4/192), 2% were infectious disease specialists (4/192), 4% were ENT (8/192), 4% were ophthalmologists (7/192), and 5% were dentists (9/192). [See Table 1]

**Table 1.**
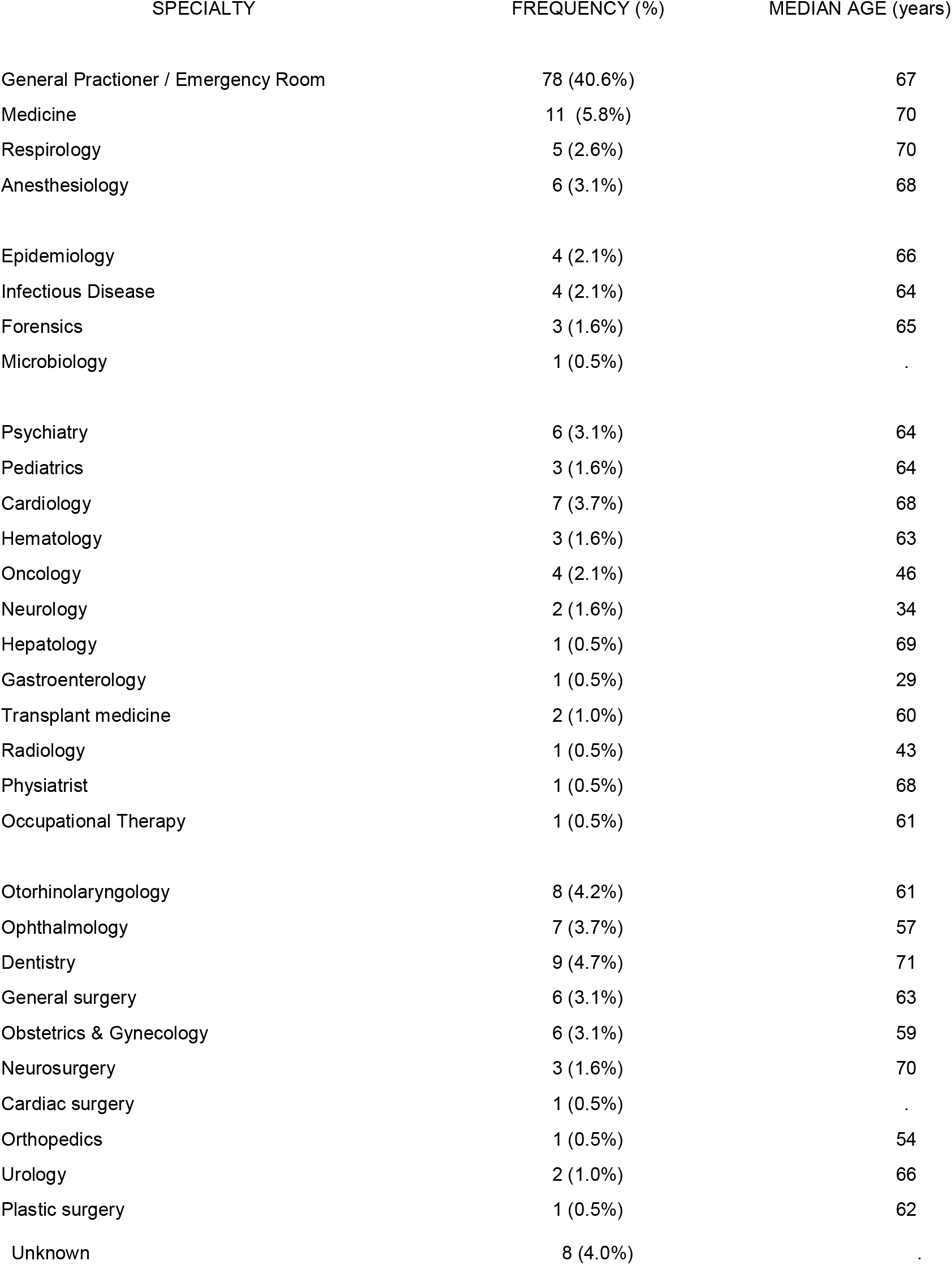
Reported Physician Deaths from COVID-19 by Practice Specialty and Median Age (n=198) on April 5, 2020

Twenty-two percent (42/191) of the physicians practiced a surgical specialty with average age 63.4 years, compared to their primary care or medicine colleagues of average age 63.6 years. There was no statistically significant difference between the ages of the surgeons and non-surgeons. (p=0.96)

The countries with the most reported physician deaths were Italy (79/198; 40%), Iran (43/198; 22%), China (16/198; 8%), Philippines (14/198; 7%), United States (9/198; 5%), Indonesia (7/198; 4%) and France (6/198; 3%). [See Table 2]

**Table 2.** Reported Physician Deaths from COVID-19 by Country (n=198) on April 5, 2020

| Country | Frequency (%) | Median Age (Years) |
| --- | --- | --- |
| Italy | 79 (39.9%) | 69 |
| Iran | 43 (21.7%) | 54 |
| China | 16 (8.1%) | 51 |
| Philippines | 14 (7.1%) | 62 |
| USA | 9 (4.6%) | 67 |
| Indonesia | 7 (3.5%) | 58 |
| France | 6 (3.0%) | 67 |
| Spain | 6 (3.0%) | 59 |
| United Kingdom | 4 (2.0%) | 66 |
| Brazil | 2 (1.0%) | 53 |
| Mexico | 2 (1.0%) | 45 |
| Turkey | 2 (1.0%) | 67 |
| Canada | 1 (0.5%) | 62 |
| Egypt | 1 (0.5%) | 50 |
| Greece | 1 (0.5%) | - |
| Honduras | 1 (0.5%) | 56 |
| Pakistan | 1 (0.5%) | 26 |
| Poland | 1 (0.5%) | - |
| Serbia | 1 (0.5%) | 59 |
| South Korea | 1 (0.5%) | 60 |

### DISCUSSION

Death in the line of duty is the doctor’s ultimate sacrifice, which may be compounded when physicians unknowingly infect family members. ^(6)^ The general public may not comprehend the importance of self-isolation measures to contain the COVID-19 pandemic until a physician dies fighting the virus. ^(7)^

We acknowledge the limitations of this paper. The number of physician deaths from COVID-19 is likely under-reported given the rapidly changing course of the pandemic, and because national mortality figures may not be available or accurately reported if all patients are not tested. Although reliance on social media searches is suboptimal, it may be the only accessible source of information, especially in countries with limited press freedom. We could not discern if the physicians died from COVID-19 or underlying conditions with associated viral infection. Pre-existing medical morbidities were not uniformly reported. The paper does not examine the number of physicians infected with COVID, who have not died; nor does it enumerate the mortality or our nursing and allied health colleagues.

Physician deaths from COVID-19 may vary between countries due to the time of disease outbreak, varying public health resources, governmental policies and controls to enforce quarantine and social distancing and mask wear, the amount of testing performed, ascertainment bias, available medical supplies and technology and social greeting habits. ^(8)^ Nations such as Singapore, South Korea and Taiwan that exercised decisive action to prevent travel from affected regions, strict enforcement of quarantines and widespread testing contained the epidemic and there are few social media reports of physician deaths. ^(9)^ South Korea had no physician or nurse deaths from COVID-19 until 10 weeks after their disease outbreak. ^(10) (11)^ As the disease onset in North America and the United Kingdom lagged behind Asia and Europe, the coming weeks may reveal a rise in COVID-19 related physician deaths in these new regions. On April 5, 2020 there was at least twelve COVID-19-related physician deaths in North America. An increasing number of North American physicians have contracted COVID-19 and are in critical care. ^(12)^

Physicians from almost all the medical specialties from psychiatry to urology have succumbed to work-related COVID-19. Physicians working in the airway such as dentists, otorhinolaryngologists, and anesthesiologists are especially at risk for COVID-19 infection and this group comprised 12% of all physician deaths. Dentists are in close proximity to oral secretions for prolonged periods and their high-speed handpiece and ultrasonic instruments aerosolize body fluids. In our review 5% of the fatalities were in dentists. However, in a recent paper from China ^(13)^ no dentists were reported to die from COVID-19 contracted during patient encounters. Ophthalmologists also work for extended periods of time close to oronasal secretions. Dr. Li Wenliang, the ophthalmologist from China who first alerted the world to COVID-19, died from the disease at the age of 33 years. Subsequently, two more ophthalmologists at Li’s workplace passed from COVID-19, perhaps due to prolonged proximity to the airway of infected patients during ophthalmoscopy, tear transmission of the virus or from nasolacrimal manipulations.

Lack of personal protective equipment (PPE) and inadequate PPE were commonly cited as a cause of death especially in developing nations and Italy. ^(14)^ When PPE is available, proper donning and doffing techniques are required. If adequate manpower is available work teams with a primary examiner, safety monitor, and scribe may decrease the risk of inadvertent contamination.

Patients who lied about their travel history of infectious contacts have also been attributed to physician deaths from COVID-19. ^(9)^ Mandatory passport inspections during pandemics might avert the former. Meticulous public health investigations and expeditious location of all contacts assisted by smartphone tracking might prevent the latter.

Purported risk factors for severe disease and death in COVID-19 include older age, male gender, hypertension, diabetes mellitus, cardiovascular disease, chronic lung disease and immunocompromise. ^(15)^ Senior physicians and those with comorbidities ideally should not be assigned to front-line work with COVID-19 patients, but instead duties such as video or phone assessments, consulting or public liason. ^(16)^

Italian physicians have suggested that a community-centred or home care system for COVID-19 rather than a hospital-focused system might decrease the transmission of disease and physician exposure. ^(17)^

Patient monitoring technologies that track pulse oximetry and vital signs and medical robots allow doctors and nurses to remotely assess and treat infected patients can enhance patient care and decrease risks to health care workers. At present robots that take temperatures, deliver foodand supplies, and disinfect rooms have been described. ^(18)^ However, in countries without resources for even gloves or masks, robots are wishful thinking.

In summary, physicians from almost all medical specialties have succumbed to COVID-19, and all require task-appropriate personal protective PPE. We are in the early phase of the pandemic and the number of physician fatalities will increase. Doctors who were 57 years of age or older accounted for three-quarters of COVID-19-related deaths. If senior or retired physicians are recalled to work during the COVID-19 crisis, ^(19)^ consideration should be made to place them away from the front line due to the increased risk of mortality, and the increased likelihood of burden to the medical system if they become infected.

## Data Availability

Data will be provided on request

## Copyright

Edsel Ing has the right to grant on behalf of all authors and does grant on behalf of all authors, an exclusive licence (or non exclusive for government employees) on a worldwide basis to the BMJ Publishing Group Ltd to permit this article (if accepted) to be published in BMJ editions and any other BMJPGL products and sublicences such use and exploit all subsidiary rights, as set out in our licence

## Transparency Declaration

Edsel Ing affirms that the manuscript is an honest, accurate, and transparent account of the study being reported; that no important aspects of the study have been omitted; and that any discrepancies from the study as planned (and, if relevant, registered) have been explained.

